# MALARIA PARASITAEMIA ASSOCIATED WITH ABO BLOOD GROUPINGS AND RHESUS FACTOR AMONG PATIENTS ATTENDING SPECIALIST HOSPITAL BAUCHI, BAUCHI STATE, NIGERIA

**DOI:** 10.1101/2025.07.23.25331896

**Authors:** Muhammad Auwal Gidado, Adamu Babayo Samaila, Muhammad Ruqayyah Hamidu

**Affiliations:** Biological Sciences Department, Abubakar Tafawa Balewa University, Bauchi State, Nigeria

**Keywords:** Malaria, blood group, rhesus factor, Specialist Hospial Bauchi

## Abstract

Malaria parasitaemia associated with ABO blood groupings and Rhesus factor associated with patients attending specialist hospital Bauchi was conducted using cross-sectional approach where 306 samples collected and both thick and thin blood film was prepared for the both male and female based on patients blood groups and Rhesus factor. The microscopic observation revealed that, out of 306 samples, 55% male is infected, while 45% females infected. Blood group A has 36.7% was highly significant than the other blood group types, while Blood group O 14.8% recorded the lowest prevalence. Out of the 306 examined, it was revealed that the 89.7% positive Rhesus factor were infected while 10.3% were negative Rhesus Factor. This study investigated the distribution of these blood group systems and assessed the association of malaria infection with the ABO blood groups among patients attending specialist hospital Bauchi. Blood specimens from venus of 306 patients were examined for malaria parasites using Field stains method. ABO and Rhesus blood group antigens tests were also performed using standard tile protocols. The prevalence of blood group A was highly significant than the other blood group types. There is no consensus association between malaria infection and ABO blood groups but the prevalence of higher malaria parasite density was significantly greater with blood group A (36.7%). In conclusion, blood group A was the most prevalent blood group in the study and Male with blood group A positive rhesus factor appeared to be more susceptible to higher level of malaria parasitemia. Males were more predisposed to *P. falciparum* infections than females. Apart from the young children and pregnant women, the government should include the old people (60 and above years) and males in the current control strategy for malaria in order to reduce malaria transmission. These individuals are considered to be the most susceptible. More awareness about blood group should be created among residents of Bauchi state as this would go a long way in reducing morbidity and mortality due to malaria and perhaps other blood diseases. Future work should consider the haemoglobin genotypes in addition to ABO blood grouping and Rhesus factor to reveal the exact picture of the association.

## INTRODUCTION

Malaria is an infection caused by haemo parasitic protozoan of the genus *Plasmodium* which is mostly transmitted to humans via bite from infected female *Anopheles* Mosquitoes (Awute *et al*., 2021). As of 2017, the global cases of malaria have reached 219 million with about 435 000 deaths, particularly in sub-Saharan Africa (WHO, 2021). The reason why the prevalence is high in sub Saharan Africa is because their weather is suitable for the parasite to multiply. For many centuries, the parasite has victimized uncountable number of people. This infection is mainly caused by multiple etiological agents which comprised of:*Plasmodium falciparum (p.falciparum), malariae, ovale, vivax* and *knowlesi* (Abdllah, 2023). *P. vivax* is responsible for the highest amount of malaria transmission worldwide, but the highest number of causalities are caused by P. *falciparum* which accounts for about 90 % of deaths from malaria and most of the victims are children below 5years old (*Akanbiet al*.,*2022*).

Blood is a body fluid in humans and other animals that delivers necessary substances such as nutrients and oxygen to the cells and transports metabolic waste products away from those same cells. In vertebrates, it is composed of blood cells suspended in blood plasma. Plasma, which constitutes 55% of blood fluid, is mostly water (92% by volume), and contains dissipated proteins, glucose, mineral ions, hormones, carbon dioxide (plasma being the main medium for excretory product transportation), and blood cells themselves. Albumin is the main protein in plasma, and it functions to regulate the colloidal osmotic pressure of blood (Igbenghu*et al*., 2022). ABO blood group system is genetically controlled and protein of various ABO groups differs significantly in different population and ethnic groups. Thus, any national or international study reporting association of ABO groups with a disease must use population frequency of ABO groups as the base for comparison (Cserti and Dzik, 2020). Almost always, an individual has the same blood group for life, but very rarely an individual’s blood type changes through addition or suppression of an antigen in infection, malignancy, or autoimmune disease. Another more common cause in blood type change is a bone marrow transplant (Pathirana *et al*., 2019).

## MATERIALS AND METHODS

### Description of the Study Area

Bauchi State occupies a total land area of 49,119 km^2^ (18,965 sq mi) representing about 5.3% of Nigeria’s total land mass and is located between latitudes 9° 3’ and 12° 3’ north and longitudes 8° 50’ and <u>11° east</u>. Bauchi State is one of the states in the northern part of Nigeria that span two distinctive vegetation zones, namely, the Sudan savannah and the Sahel savannah. The Sudan savannah type of vegetation covers the Southern part of the state. Here, the vegetation gets richer and richer towards the South, especially along water sources or rivers, but generally the vegetation is less uniform and grasses are shorter than what grows even farther South, that is, in the forest zone of the middle belt.

The Sahel type of savannah, also known as semi-desert vegetation, becomes manifest from the middle of the state as one moves from the state’s south to its north. This type of vegetation comprises isolated stands of thorny shrubs.

Consequently, rains start earlier in the southern part of the state, where rain is heaviest and lasts longer. Here the rains start in April with the highest record amount of 1,300 millimeters (51 in) per annum. In contrast, the northern part of the state receives the rains late, usually around June or July, and records the highest amount of 700 millimeters (28 in) per annum.

### Sample collection

Blood samples were collected from the patients. The patient’s name, date and time of collection were labeled on the EDTA-anti coagulant tubes and pre-cleaned microscope glass slides. The site for blood collection was cleaned well using 70% methanol then allowed to air dry. About 3 ml of venous blood from the patient was collected into an EDTA-anti coagulant tube using a 5 ml syringe and needle set and examined in the laboratory.

### Data Analysis

Data obtained, Comparison of levels of parasitaemia amongst individuals of different ABO and Rhesus blood groups was statistically analyzed using Chi-square (χ^2^) on SPSS computer software version 22 of 2021.

### Results

Out of the total of 306 blood samples collected, 191 (62.4%) were found to be infected with Malaria infection. The prevalence was found to be highest among under five children (0 10). Similarly, the prevalence was higher among males 100 (52.4%) than female 91 (47.6%). Majority of the patients were rhesus positive 121(63.4%) while 70(36.6%) rhesus negative. High percentage of blood group A 59 (30.4%) was observed among the study participants. There was highly significant difference between the various ABO Blood group and prevalence of malaria infection by age of subjects (P>0.05).

**Table 1.**
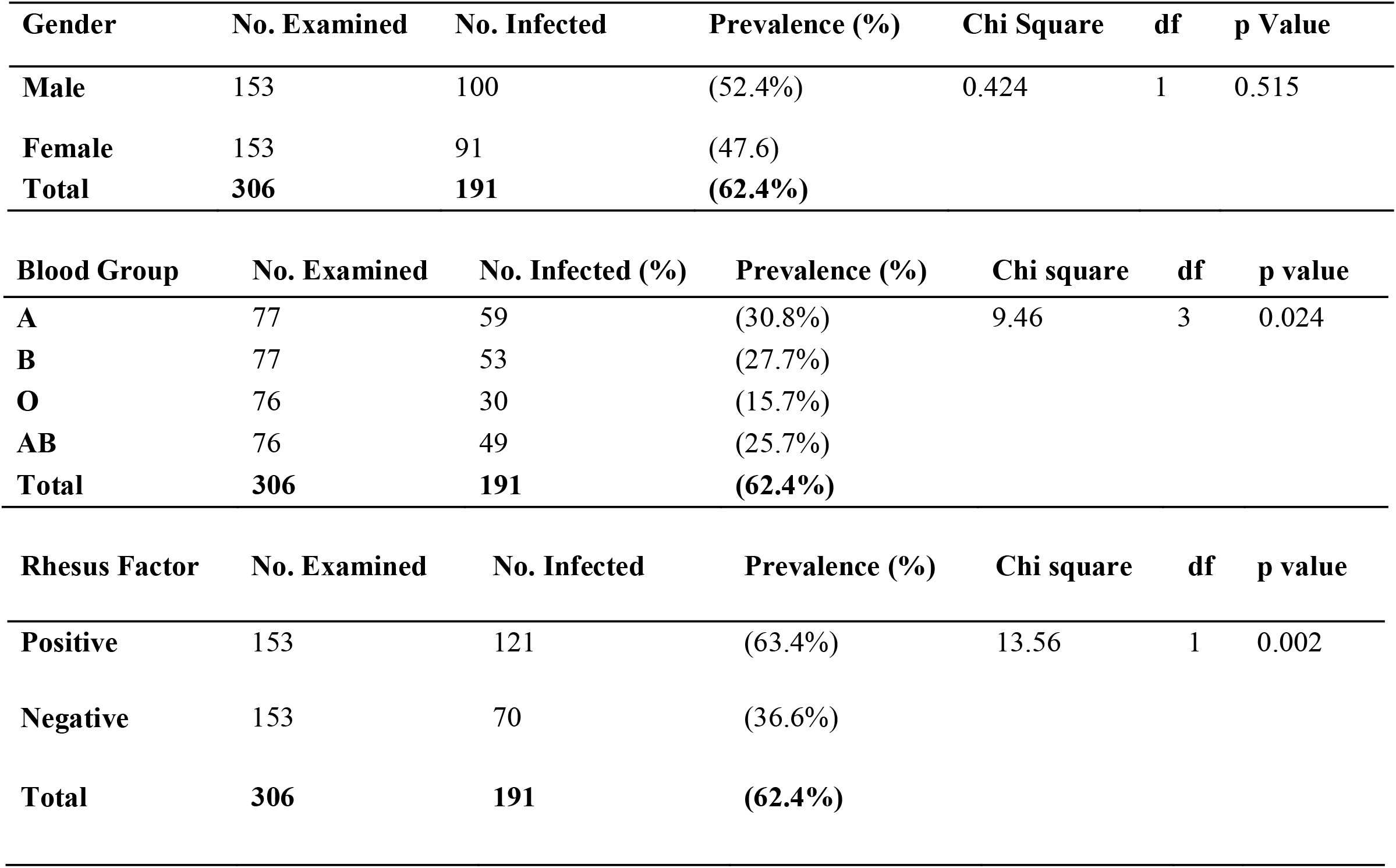
Overall Prevalence of Malaria by gender, blood group and Rhesus factor.

All ABO blood groups showed the presence of malaria infection to a certain level, 59(30.4%), 53(27.7%), 49(25.7%) and 30(15.4%) for A, B, AB, and O respectively with significant difference P<0.05). Male were more prone to maria infection than female accounting 100 (52.4%) reason for this has not been established scientifically but may be due to the fact that males within the study area are the most exposed to mosquitoes biting than females while women in the study area are mostly not exposed due to religion and cultural believes.]. Apart from exposure, stress (physically and mentally) due to their responsibility, may also be the predisposing factor, however some suggested that genetic factors could play a role by endorsing female with immuno regulatory potentials to cope better with some disease. This is also in agreement with studies by Agbonlahor *et al*.,2019.

**Table 2.** Age Distribution of Patients attending Specialist Hospital Bauchi.

| Age Group | No. ined | No. Infected (%) | Prevalence (%) | chi | df | p value |
| --- | --- | --- | --- | --- | --- | --- |
| 0–9 | 44 | 40 | (20.9%) | 28.5 | 6 | 0.001 |
| 10–19 | 44 | 25 | (13.1%) |  |  |  |
| 20–29 | 44 | 23 | (12.0%) |  |  |  |
| 30–39 | 44 | 15 | (7.9%) |  |  |  |
| 40–49 | 44 | 19 | (10.0%) |  |  |  |
| 50–59 | 43 | 31 | (16.2%) |  |  |  |
| 60–69 | 43 | 38 | (19.9%) |  |  |  |
| **Total** | **306** | **191** | **(62.4%)** |  |  |  |

**Table 3.**
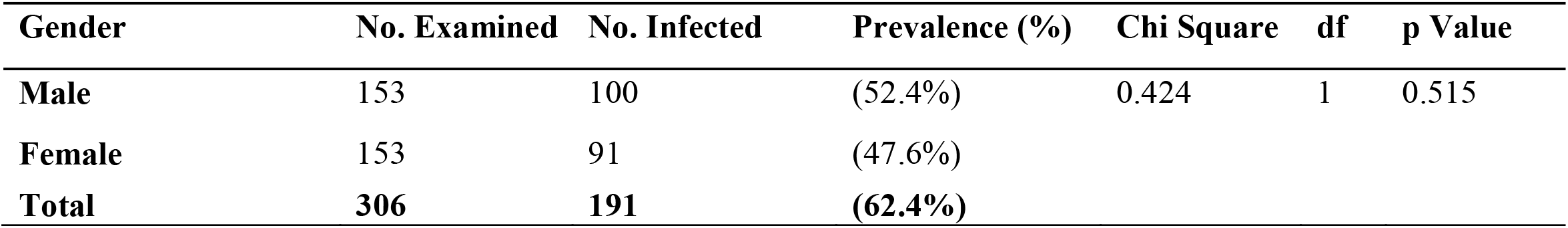
Association Between Gender and Malaria Infection Status.

**Table 4.**
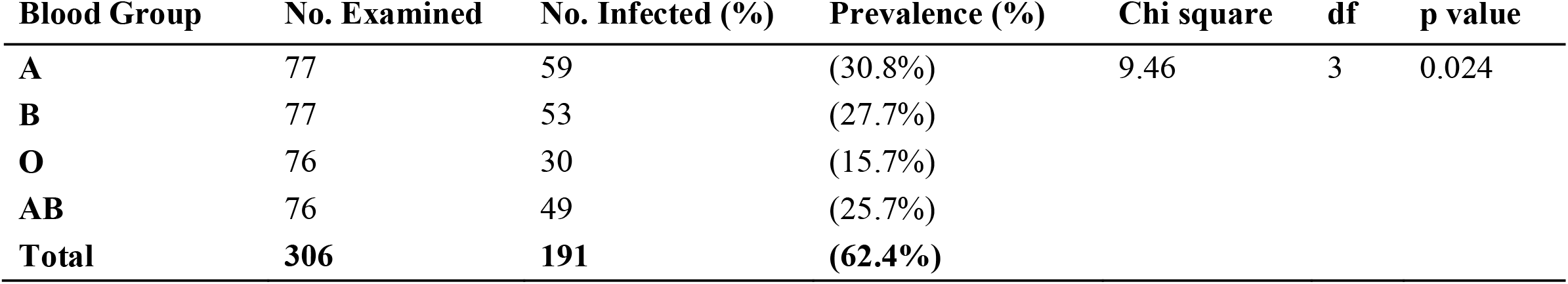
Association Between ABO Blood Groups and Malaria Infection Prevalence Blood Group No. Examined No. Infected (%) Prevalence (%) Chi square.

**Table 5.**
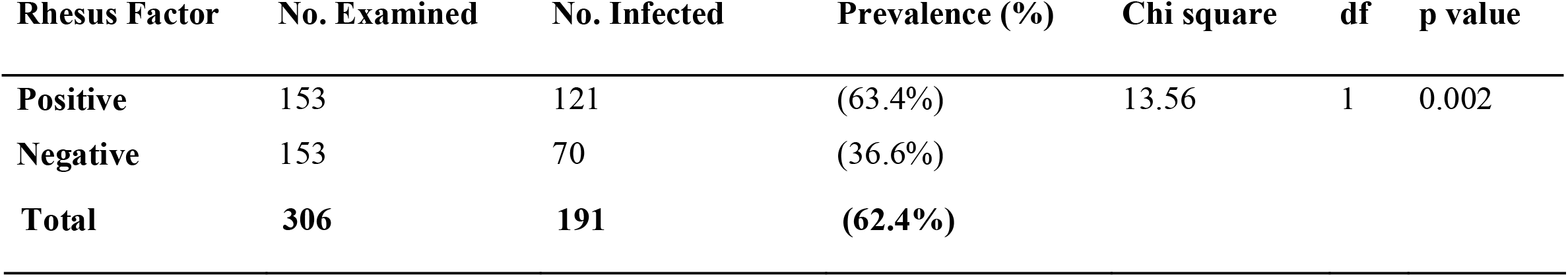
Association Between Rhesus Factor and Malaria Infection Rhesus Factor No. Examined No. Infected Prevalence (%)

Moreover, out of 306 patients that were examined only 70 (36.6%) patients were Rhesus negative, the remaining 121 (63.4%) patients were Rhesus positive, this is in agreement to previous findings. Individuals with blood group A were found to be more prone to malaria infection (66.3%) compared with other Blood groups. This could be as a result of the fact that both ABO and Rh blood group have attracted enormous attention regarding their association with genetic and infectious diseases, previous studies on patients of cancer and tumor, heart disease, and parasitic and viral infections indicated associations of ABO and Rh blood groups. Moreover, Malaria parasites are more common and severe in group A individuals compared with other Blood groups. Blood group O, B, and AB has their corresponding antigens whereas A has none. Malaria parasite find it hard to invade the red cells of individuals with the O, B and AB groups and required to digest the surface Antigen through enzymatic activity. There is however evidence that other Blood groups were almost at the same level of morbidity, and thus there is need for assessment of relation between ABO and Malaria severity. Wolofsky et al.2022, showed that there was no significant relationship between the prevalence of malaria and ABO blood groups and *P. falciparum* sporozoites invade and mature irrespective of the different ABO blood groups.

## CONCLUSION

Males were more predisposed to malaria infections (52.4%) than females (47.6%) among patients attending Specialist Hospital Bauchi. Malaria prevalence level in patients exermined Specialist Hospital Bauchi were 59(30.4%), 53(27.7%), 49(25.7%) and 30(15.4%) for A, B, AB, and O respectively with significant difference P<0.05). Blood group O confer a certain degree of protection against severe courses of malaria. This is evidenced in that O+ individuals had the least mean parasitaemia levels in which when it was subjected to Chi-square test, it showed lack of association with malaria infection

## RECOMMENDATION

Apart from the young children and pregnant women, old people (≥60 years) should be included in the current control strategy for malaria in order to reduce malaria transmission. These individuals were found to be the most susceptible group in the study Area. People with blood Group A+ should wear cloths that will cover their entire body, and they should stay in a close door with mosquitoes nets as they are more prone to malaria infection.

Those with negative positive rhesus factor should take proactive measures against malaria, such as taking prophylaxis drugs

## Data Availability

All data produced in the present work are contained in the manuscript

## CONSENT AND ETHICAL APPROVAL

Ethical approval for this study was obtained from the **Bauchi State Health Research Ethics Committee (HREC)**.located at the Ministry of Health, Bauchi State, Nigeria. The committee reviewed the study protocol and granted ethical clearance (Approval Reference No: HREC/ADM/MLS/0973, dated 22/01/2025.

In addition, permission to conduct the study was also obtained from **Bauchi State Specialist Hospital Management Board**, which provided institutional support and access to the study site.

The ethics committee confirmed that the study meets all ethical standards and approved the research. All procedures performed in the study involving Human participants were in accordance with the ethical standards of the institutional and national research committee.

